# “It does help but there’s a limit…”: Young people’s perspectives on policies that restrict hot food takeaways opening near schools

**DOI:** 10.1101/2024.07.17.24310555

**Authors:** B Savory, C Thompson, S Hassan, J Adams, B Amies-Cull, M Chang, D Derbyshire, M Keeble, B Liu, A Medina-Lara, OT Mytton, J Rahilly, N Rogers, R Smith, M White, T Burgoine, S Cummins

## Abstract

**Background:** Local authorities (LAs) in England are increasingly using the planning system to control the proliferation of hot food takeaway outlets (‘takeaways’) near schools as part of a range of policies to promote healthy weight in children. These ‘takeaway management zones’ (TMZs) include restrictions on planning permission to open new takeaways within a certain distance of a school. In this qualitative study we explore young people’s perspectives of TMZs.

**Methods:** We purposively recruited 46 young people (aged 11-18 years old) attending secondary school across two contrasting London LAs with operating TMZs. We conducted semi-structured, walking group interviews (“go-alongs”) in January-February 2023 in the local food environment close to participants’ schools. We analysed data using framework analysis.

**Results:** Participants generally viewed TMZs as reasonable and uncontroversial but were not always aware that TMZs were in operation. Although participants understood that TMZs prevented new outlets from opening, they observed they did not seem to reduce existing provision. This was viewed positively as it did not result in the closure of local takeaways perceived as important components of the social fabric of school life. Participants believed that the potential health impact of TMZs is limited by their exclusive focus on takeaways as other food retail commonly patronised by young people, such as convenience stores, are important sources of unhealthy food. Participants also identified inadequacies in the wider food environment, including the school dining environment and access to food delivery apps.

**Conclusions:** Our findings suggest that although young people find TMZs acceptable and believe they have some positive impact on diet, they did not perceive TMZs as effective as they could be. Participants articulated that the management of takeaways on their own is unlikely to reduce exposure to unhealthy foods. Widening the remit of planning policy to include outlets selling convenience foods may be important for policy optimisation.

**Highlights:**

- Young people’s perspectives of takeaway management zones are explored
- Acceptability of zones was high as the existing food environment remains unchanged
- The impact of takeaway management zones was deemed limited by its focus and scope
- Considering wider aspects of the food environment is key for policy development
- Future policy should also consider young people’s social and emotional needs

## Introduction

The environments where we source and consume food can influence dietary choices in a way that is detrimental to health (Neve & Isaacs, 2022; WHO, 2016). The association between exposure to a poor-quality food environment and diet is well known and may contribute to the generation and maintenance of diet-related inequalities in health (Hallum et al., 2020; NHS Digital, 2019). In response, environmental interventions have been proposed as components of local and national public health strategies to improve population diet and reduce dietary inequalities (Penney et al., 2014; Public Health England, 2019).

As adolescents spend 40% of their waking time in school each week, the food environment in and around schools may significantly influence their dietary choices and behaviours (França et al., 2022; Gonçalves et al., 2021; Ohri-Vachaspati et al., 2023). Evidence suggests that adolescents are frequent consumers of hot food takeaways (‘takeaways’ hereafter) with purchases occurring during school breaks, lunch times and on their journey to and from school (Macdiarmid et al., 2015; Taher et al., 2019; Thompson et al., 2018). One study on UK secondary school students (aged 11-14) found that more than half purchased fast food or takeaways at least twice a week, and 1 in 10 everyday (Patterson et al., 2012).

Foods sold in takeaways are typically energy-dense, nutrient-poor, and served in larger portion sizes (Jaworowska, 2014; Keeble et al., 2019b). Regular consumption of takeaways is associated with poor health outcomes in adults and children (Duffey et al., 2007; Penney et al., 2017; Pereira et al., 2005) and evidence suggests some association between physical exposure to takeaways and excess weight (Burgoine et al., 2016; Jiang et al., 2023; Patterson et al., 2012). Obesity in adolescence has been identified as a strong predictor of adult obesity, with interventions targeting dietary behaviours during this developmental stage proving particularly effective for weight reduction (Neufeld et al., 2022; Simmonds et al., 2016).

Interventions targeting the food environment around schools have been shown to impact the dietary behaviours of young people and the wider population (Cotton et al., 2023; Ohri-Vachaspati et al., 2023). One such intervention is the development of planning policy to manage applications for new takeaway outlets (Keeble et al., 2019b). In 2019 half of England’s 325 LAs had developed takeaway planning policies, with the most common approach involving implementing takeaway ‘exclusion zones’ around schools where planning permission for new takeaway outlets may be denied or restricted (Keeble et al., 2019b). Although the term takeaway ‘exclusion zone’ is commonly used by LAs, in this paper we use the term ‘takeaway management zones’ (TMZs) to capture the varied approaches adopted by different LAs. By 2019, 41 LAs had adopted TMZs (Keeble et al., 2019b). Existing takeaway outlets that fall within the TMZ remain unaffected by the policy and the size (e.g. 400m or 800m), shape (the point from which the exclusion zone starts) and inclusion criteria (primary and secondary schools or just secondary) vary across LAs. The most common specification is a 400m TMZ, which is believed to equate to a 5-minute walk from the entrance of a school (Homes and Communities Agency, 2006).

Despite the focus on schools, there has been limited research investigating young people’s perceptions of these interventions. Given that public attitudes often impact the effectiveness of public health policy, exploring the views of the target population has the potential to increase policy acceptance and impact (Diepeveen et al., 2013). Public involvement in policy-making processes can further increase acceptance, and the value of youth participation in the creation and development of policies that affect them is increasingly being recognised (Macauley et al., 2022; Patton et al., 2016). We therefore aimed to explore the acceptability, perceived effectiveness, and barriers and facilitators of the policy’s impact amongst young people (aged 11-18) attending secondary schools in LAs with TMZs in operation.

## Methods

### Setting

Data were collected in the London Boroughs of Islington and Redbridge in February and March 2023, and were selected with the aim of including pupils from diverse socioeconomic backgrounds. London boroughs have some of the highest densities of takeaways in England (Keeble et al., 2019a).

Islington is the second most densely populated LA in London and one of the most deprived (Greater London Authority, 2023; Trust for London, 2023a). In contrast, Redbridge is less densely populated and deprivation is average compared to all London Boroughs (Greater London Authority, 2023; Trust for London, 2023b). In 2013 and 2018 respectively, Islington and Redbridge councils introduced policies to manage the proliferation and concentration of takeaway outlets. Islington council TMZ policies specify that planning permission for new takeaway outlets within a 200m radius of primary and secondary schools should be resisted (Islington Council, 2016) and Redbridge, new takeaway outlets within a 400m radius (Redbridge Council, 2018).

### Sampling and recruitment

A purposive sample of 38 state-funded secondary schools across Islington and Redbridge were initially approached for recruitment. All secondary schools within selected LAs were contacted twice and provided with study information sheets and flyers to aid recruitment, with a total of four schools (two schools in each LA) agreeing to facilitate and help supervise go-along group interviews. School contacts were asked to recruit up to 9 students per go-along group interview, preferably with a mix of ages and genders (if coeducational). We conducted two go-along interviews in schools where more than students signed up to participate.

A total of 46 participants aged between 11-18 years were recruited. Participant and school characteristics are summarised in Table 1. As the school experience varies by age, participants were split into age groups of younger participants, aged 11-15 years, and older participants, aged 16-18 years.

**Table 1:** Go-along interview participant characteristics.

| Local Authority | School | Description of school | Interview number/s | Age Range of interview participants (years) | Female Participants n=35 | Male Participants n=11 | Total Participants n=46 |
| --- | --- | --- | --- | --- | --- | --- | --- |
| Redbridge | A | Single-sex, state | 1 | 16-18 | 8 | 0 | 8 |
|  |  |  | 2 | 16-18 | 7 | 0 | 7 |
| Redbridge | B | Co-ed, state | 3 | 11-14 | 4 | 5 | 9 |
|  |  |  | 4 | 16-17 | 5 | 2 | 7 |
| Islington | A | Co-ed, state | 5 | 16-17 | 3 | 4 | 7 |
| Islington | B | Single-sex, state | 6 | 12-13 | 8 | 0 | 8 |

Amongst participants aged 11-15 years, informed consent was obtained from both parents/guardians and participants themselves. Amongst older participants, only participants provided informed consent. Full ethical approval was obtained from the London School of Hygiene and Tropical Medicine Research Ethics Committee (Ref/ 26337).

### Data Collection

Given this study’s aim to understand young people’s perceptions and experiences of the food environment around their school, go-along interviews were chosen. This mobile method allowed reflections on the everyday food practices of participants as they unfolded in time and space (Carpiano, 2009; Kusenbach, 2003). Previous studies have found that participatory approaches with children and young people, such as go-along interviews, have the potential to reduce the power imbalance between researcher and participants by situating the young person as the expert guide (Hayball & Pawlowski, 2018; Horgan et al., 2022).

Interviews were conducted following a semi-structured topic guide designed and piloted initially in consultation with PPI groups at the University of Hertfordshire (including adolescents) and was developed iteratively throughout data collection. Questions and prompts aided investigation into policy acceptability, its perceived impacts, and barriers to effectiveness. Each interview began at the school site with an introduction to the study and continued on foot around the local food environment. The route chosen by participants typically involved walking to the nearest high street with takeaway and non-hot food outlet outlets that they identified as most popular with school pupils. Figure 1 illustrates an example go-along interview route.

**Figure 1:**
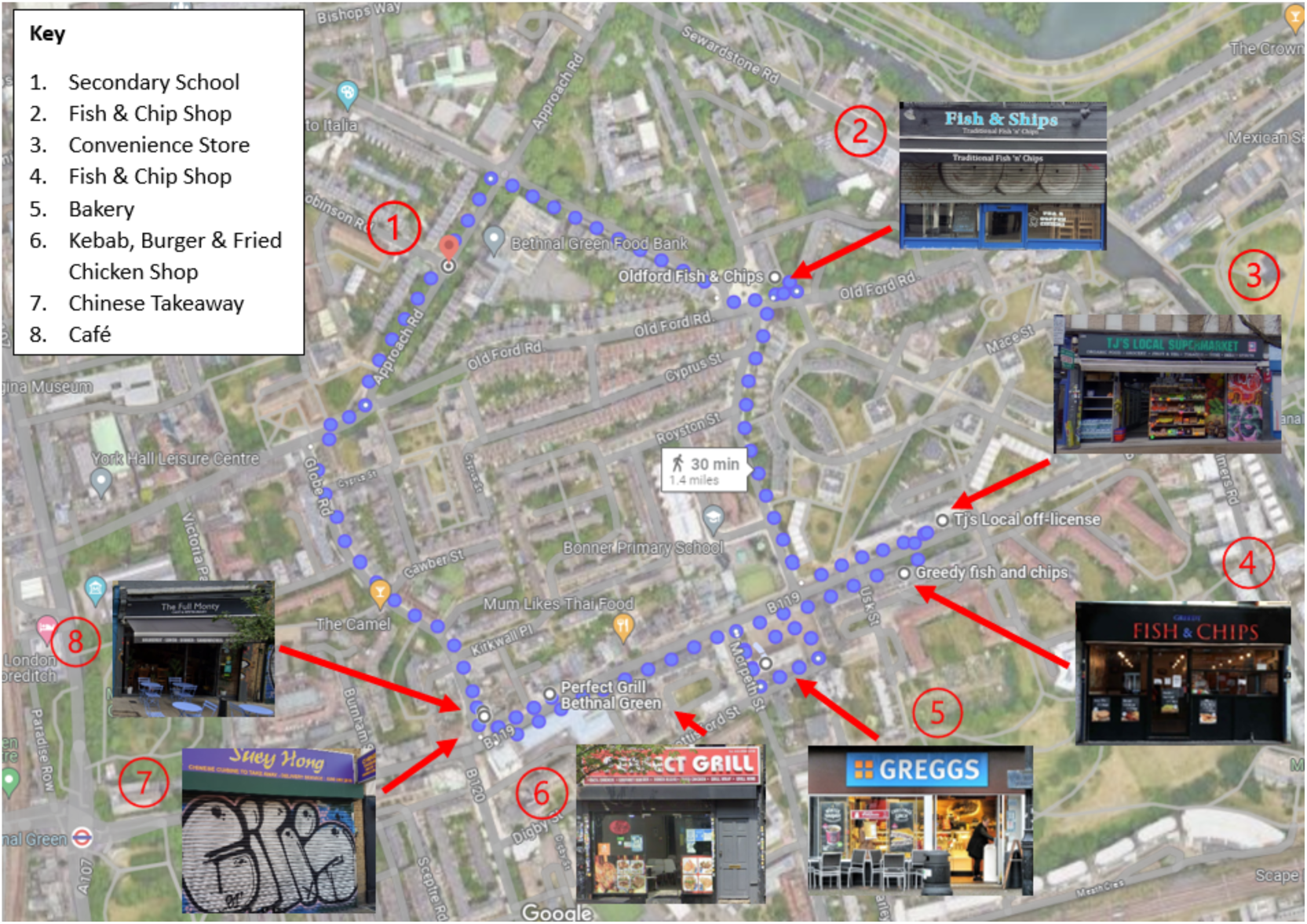
An example go-along interview route, starting at a defunct secondary school in London and continuing on foot around the local food environment.

Six go-along interviews with 46 participants were conducted, led by BS, and lasted an average of 42 minutes. Interviews were audio-recorded and notes and photos were taken to aid analysis and contextualise the local food environment. Prior to interview, participant background questionnaires were completed to collect data on personal characteristics (age, ethnicity, school, home), takeaway consumption, and purchasing behaviour. Trips were conducted during school hours and were accompanied by a school chaperone; all participants were provided with a £20 shopping voucher to compensate them for their time.

### Data analysis and reflexivity

Audio-recordings were transcribed verbatim by an external company and checked manually by BS; all identifying information was removed and all participants were pseudonymised. Transcripts were imported into NVivo 12 software for coding and data was analysed using Framework Analysis (Ritchie & Spencer, 1994). Framework Analysis is particularly suited to applied public health research because it allows for specific research questions and aims to be addressed through a flexible and comparative analytical approach. Data are systematically organised into deductively derived themes driven by *a priori* research questions and then combined to establish an inductively derived framework, which remains fluid and open to interpretation. Throughout the data analysis process, discussions were held with SC and CT to ensure a range of perspectives were considered when interpreting the findings. Consensus on the framework was reached at the mapping and interpretation stage of analysis.

The researchers acknowledge that the lead author’s background and positionality may have influenced the creation and interpretation of the data. BS is a young, White woman with 3 years’ previous experience working in a London secondary school, giving her a unique insight into the experiences of UK school pupils. This background provides valuable context but also necessitated careful consideration of how her potential biases and preconceptions may have shaped the research; BS remained critically aware of these factors throughout the research process.

## Results

In the following sections, we explore participant experiences of the food environment in and around their schools, paying particular attention to perspectives concerning TMZs. Our analysis identified three main themes: awareness and acceptability of TMZs; perceived impacts; perceived barriers to effectiveness. Given the heterogeneity of findings within the last theme we divide this section into three subthemes: inadequate school food and school environment; purchasing convenience food and drinks from non-hot food outlets; food delivery apps.

### Awareness and acceptability of TMZs: “I don’t think I would notice”

Participants found TMZs broadly acceptable as they were unaware that it was in operation. Rather than threatening the existing, and sometimes much-loved, local outlets, the policy’s focus on restricting the growth of new takeaways meant that its potential effects were not directly observable. While the policy may have restricted the opening of new outlets near their school, participants were generally satisfied with the current food retail offer and were loyal to existing outlets. Some participants expressed that their contentment with the current food options available influenced their receptiveness to TMZs:

> *“I don’t think I would notice if new takeaways weren’t allowed to be made anymore. I don’t think I would notice ‘cause I’m content with the options we have here…but for as long as we don’t have it, we wouldn’t know the possibility…I think I’m fine without.”*

> (Pupil from Redbridge A1, aged 16-18)

When visiting local food outlets, participants were keen to highlight their social, cultural and economic value. Participants spoke fondly of the enduring relationships between shop workers and the local school pupils, considering them fundamental features of the school community and its culture. The personalised service provided by such retailers was also frequently raised, with participants speaking of special treatment in the form of student discounts and deals, like *“buy two, you get one free”* (Pupil from Redbridge A1, aged 16-18). Participants, including the following, exhibited a strong sense of loyalty towards local businesses:

> *“They’re [the local Fish & Chip shop] like very, acquainted with tradition and that place has been around for, like, so long, so it would be so weird if that chip shop just disappeared because it’s an independent one. It’s known to be in this area, so that makes it,….we have…a great attachment to it and we don’t want to get rid of it.”*

> (Pupil from Redbridge A2, aged 16-18)

Takeaway outlets (including those with seating and the option to eat in) were referred to as important community spaces for socialisation, and their patronage a marker of social belonging and inclusion. The acts of socialising and eating takeaway food seemed to be intrinsically linked for participants; it was routine to do both things at once. Some participants specifically raised the issue of takeaways filling a gap in alternative spaces for young people to meet and “*socialise*” after school and on the weekends. One participant described the lack of *“accessible”, “safe zones”* in his local community where young people can *“go to, not to order food, but just to hang out”* (Pupil from Redbridge B3, aged 11-14). In this way, these spaces transcended their function of fulfilling gustatory needs to become hubs for social interaction and passing time. Although participants often described eating takeaway food as the primary motivator for patronising takeaways, their social potential evidently played an important role in their food choices and behaviours:

> *“A lot of people go out to eat because it’s the only thing you can do around here, like to hang out.”*

> (Pupil from Redbridge A1, aged 16-18)

> *“Takeaways and fast food restaurants in the area are a way for a lot of people to socialise and by taking that away it takes away a lot of social activity and then everything’s committed to just school and you won’t be able to make those connections and relationships outside of school.”*

> (Pupil from Redbridge B4, aged 16-17)

### Perceived impacts of TMZs: “I don’t think it would make much of a difference”

Participants generally perceived TMZs to have had little tangible impact in reducing young people’s physical access to takeaways as visible concrete ‘change’ in the food environment was not observable. As a result, a policy focus on reducing takeaway growth was expressed as synonymous with a lack of significant perceived change (either positive or negative) in youth dietary behaviours.

The local context in which the policy was applied had a bearing on how participants theorised its possible impacts. In areas with numerous takeaways and other food outlets selling unhealthy foods, participants generally believed that preventing further proliferation is unlikely to achieve much as these areas are already saturated. This issue was particularly salient to the participants in this study, who all attended schools situated near a wide array of food outlets. Some participants expressed a desire to consume *“healthy”* takeaway and convenience food but struggled to access outlets near their schools selling affordable healthier options.

In the following excerpt, one participant reflects on the issues of implementing the policy in areas already highly saturated with takeaway outlets. It was explained that small reductions in the number of takeaways are unlikely to be impactful without closing existing outlets:

> *“It won’t affect that much, it might affect a lower majority of the areas. But if in places that don’t already have fast food places that would make sense, but somewhere like here, we are so close to a town centre, it won’t do much.”*

> (Pupil from Redbridge B4, aged 16-17)

While participants acknowledged that the policy may *“help”* to *“limit students’ choices”* by reducing access to takeaways near schools, they also noted the limitations of the scope and size of the TMZ itself. Participants pointed out that the policy may not apply to other areas they frequent, including places they pass through on their journeys home. For older participants typically able to *“go anywhere, anywhere you want”* (Pupil from Redbridge B4, aged 16-17) during break and lunch times and study periods, a TMZ was also not perceived as a barrier to accessing takeaway (and convenience) food during the school day. There was a strong consensus across age groups that students would willingly travel beyond the TMZ if their gustatory and social needs were not being met:

> *“I don’t think completely closing down or not closing down, but preventing new takeaway shops near schools is going to change anything ‘cause students can still travel to other areas to get food. Especially after school, which is when most people would get food.”*

> (Pupil from Redbridge A1, aged 16-18)

> *“I think it also wouldn’t fix the problem. People would just go elsewhere and try and find more takeaways.”*

> (Pupil from Redbridge A2, aged 16-18)

### Perceived barriers to effectiveness: “I don’t think it’s takeaways”

When asked about their perceptions of the policy, participants were particularly keen to discuss their beliefs about potential barriers to effectiveness. Conversations revolved around issues in the wider food environment in and outside of school, such as school food, non-hot food outlets and food delivery apps (FDAs), and their collective influence on youth dietary behaviours. Participants expressed the belief that TMZs can only achieve a partial management of the food environment, which reduces its potential to decrease access to unhealthy foods and improve diets.

### Inadequate school food and school environment

Many participants argued that the policy’s impact is compromised by the unappealing and inaccessible state of school food and school dining environments. They argued that the comparative affordability and expediency of takeaway food rendered the external food environment particularly appealing and, at points, necessary to compensate for the shortcomings of the school food environment. This juxtaposition led to a common perception among participants that improving the taste and material conditions of school food would deter young people from patronising external food outlets at break and lunch times (if permitted) and after school:

> *“I think if they improved school dinners it might actually help because a lot of people might be like, I don’t want to eat the school dinners, I’m just going to eat something after school instead from a takeaway. But if there were nicer school dinners that people really wanted to eat, then they probably wouldn’t be hungry after school, so they wouldn’t really want to go to the takeaways.”*

> (Pupil from Islington B6, aged 12-13)

The most common recommendation made for improvement related to the temporal organisation of lunchtime, which disincentivised participants from eating and socializing with their peers in school. Accounts of eating in “*noisy*” and *“busy”* canteens, often with *“long”* and time-consuming queues, were frequently contrasted with experiences of eating *“quick”* takeaways in *“open areas”*. While older participants could avoid this issue by eating outside of school, younger participants recounted instances where *“you might not have time to eat in because the queues are so long”* (Pupil from Islington B6, aged 12-13). Participants generally also shared the view that school food represented poor value for money, particularly in comparison to takeaways:

> *“It costs £2.50 to get chips, and it costs around that same amount of money to get like food from school. So if it’s going to cost the same amount, but you can get like a big portion of chips that’s more filling, and that’s warm, and faster, a lot of students might prefer to get that.”*

> (Pupil from Redbridge A1, aged 16-18)

Considering these frustrations, participants particularly objected to interventions in the external food environment, where they spend their *“own time”*, when they perceived a lack of similar government intervention in schools. There seemed to be a sense that it would be more impactful, and more acceptable, for interventions to focus on improving food provisioning in educational establishments where autonomy is ordinarily (and knowingly) restricted:

> *“Making school food healthier and more inclusive or have wider variety because then, students can eat something that they enjoy, and for not too much money, and it’ll be better for the health…the only reason students will pick unhealthy food opposed to a full meal is because it’s much more affordable, right?”*

> (Pupil from Redbridge A1, aged 16-18)

### Purchasing convenience food and drinks from non-hot food outlets

In addition to concerns around the shortcomings of the internal school food environment, most participants expressed the belief that the policy does not address crucial parts of the external food environment contributing to poor diets. Participants argued that young people are much more likely to purchase food and drinks (primarily confectionery, crisps, and sugar-sweetened beverages (SSBs)) before and after school than takeaway food. Participants found these items more affordable and socially acceptable and perceived them as readily available at all times of the day in the food environments they spent time in. As a result, they tended to believe that the policy also needs to tackle convenience stores selling an abundance of unhealthy snacks. Some participants explained that even if the policy resulted in fewer takeaways near their school, convenience stores would *“make up for”* any reduction in access to unhealthy foods:

> *“…what we … don’t make up in… fast food shops, we have so many sweet shops in this area, which kind of makes up for it… it’s not really that big of a deal that we’ve only got like one popular chip shop because everyone just goes to the sweet shop because we’ve got two just down there…”*

> (Pupil from Redbridge A2, aged 16-18)

Takeaways were generally regarded as a *“rare” “treat”* acquired *“occasionally”* after school, and convenience food and drinks were seen as *“more often”* everyday purchases. This was attributed to the relative low cost and accessibility of the latter; the foods on offer tended to be perceived as greater value for money than takeaways. Participants frequently referred to purchasing multiple *“little bits and bobs”* as a *“snack”* at a time, particularly *“cheap sweets”* (confectionery and chocolates) that could be shared with friends:

> *“I think it’s easier to go and buy sweets and, because of that, we do it more often, whereas the takeaway is more rare in that sense, in terms of the time that you have.”*

> (Pupil from Redbridge A2, aged 16-18)

Participants also reported being tempted by product marketing and the associated social value of displaying ‘trendy’ products in the school environment. In contrast to the generic *“chicken and chips”* or *“pizza”* sold in the local independent takeaways typically and served in unbranded or lesser-known packaging, convenience stores were characterized as selling branded sweets such as *“Twangers”* and *“Push Pops”* or often SSBs and energy drinks such as *“Boost”*, *“Lucozade”* and *“Mogu Mogu”*. Such product names seemed to be generally easily recognised amongst participants, often leading to animated discussions around their popularity and social value in school. Participants identified branded food and drink products as popular and valuable amongst young people for a variety of reasons, including their association with specific marketing to young people on social media or their perceived ability to increase productivity.

As most participants referred to specific school policies around bringing *“junk food”* (takeaways and convenience food and drinks) onto site, convenience or non-hot food and drinks, generally with less overt physical and olfactory features, were lauded for their relative transportability and ease at which they could be consumed surreptitiously in class:

> *“I guess if it’s sweets or crisps or something, you can put in your pocket, whereas you probably wouldn’t do that with chips.”*

> (Pupil from Redbridge A2, aged 16-18)

Across all participant groups, food and drinks purchased from convenience stores appeared to be associated more with everyday life than foods from takeaways. Younger participants (generally prohibited from leaving school at break and lunch times) pointed out that this was partly because *“the sweet shops open early”* allowing students to “*buy stuff before school…and then…also buy stuff after school”*. However, discussions surrounding takeaway consumption revealed that this was mainly because, regardless of age, participants found the food and drinks sold in convenience stores and newsagents more appealing, affordable, and socially acceptable:

> *“Sometimes if the bus is taking long, I’ll get some sweets from the shop and just enjoy it while I’m waiting for the bus. And on the way back to home, I do that as well, but sometimes I just get chicken and chips. So I wouldn’t get chicken and chips in the morning, but only after school.”*

> (Pupil from Redbridge B3, aged 11-14)

> *“We obviously don’t have like a lot of money to like buy a whole meal, so we’ll probably just get something cheap. Like a lot of people will just get some like sweets from the corner shop.”*

> (Pupil from Redbridge A1, aged 16-18)

### Food delivery apps

Participants identified food delivery apps (FDAs) as elements of a rapidly changing food environment, both in and outside of school. While FDAs were not reported to be widely used by young people, participants seemed to anticipate and predict that they would present more of a problem in the future by establishing a new opportunity to access takeaways. Links were made between the rise of FDAs and increased smartphone and debit card ownership, particularly by older participants with greater financial autonomy: *“it’s [name of food delivery app] on our phone, we can access it* whenever” (Pupil from Islington A5, aged 16-17). For older participants, generally permitted access to their smartphones during the school day, FDAs were described as having the power to collapse both the physical boundaries of the TMZ and the temporal boundaries of a short school lunchtime. FDAs facilitated access to a range of takeaways ordinarily inaccessible on foot:

> *“So I guess, stopping takeaways would help, as in, from popping up around schools. But then now with [name of food delivery app], how helpful is that going to be? So it’s difficult.”*

> (Pupil from Islington A5, aged 16-17)

> *“I don’t think it [TMZs] would help because people still just go home and order off [name of food delivery app 1], [name of food delivery app 2] things like that.”*

> (Pupil from Redbridge B3, aged 11-14)

Some participants specifically raised the issue of students using FDAs to order takeaways to the school site, particularly as *“a group of friends”* for *“celebrations”* like *“the last day”* of term. Participants expressed uncertainty around school policies on ordering food to school and reported varying levels of understanding about the potential consequences of breaching such rules. In one interview, a chaperoning teacher reinforced the ambiguity surrounding this issue, stating that students are currently able to *“order a takeaway and meet them [the delivery driver] outside of the school site”* since it is not *“technically”* the school’s “business” (Teacher from Redbridge A1) if the food is not delivered to the school reception. Participants described navigating this liminal space and ordering food *“secretively”* even when *“it’s not allowed”*.

Participants expressed similar uncertainty over different FDAs’ policies on delivering to school sites. While some reported instances in which FDAs generally refused delivery to their school, others spoke of individual drivers delivering orders *“secretly”*, often to just outside school gates. There was consensus amongst participants that FDAs remain largely unregulated in both the in-school and out-of-school environment and that any existing rules could be bypassed:

> *“Yes some people can order and provide, to be honest it depends on what delivery driver you get, some will just plainly just say no, but some will for the money, they’ll come secretly.”*

> (Pupil from Redbridge B3, aged 11-14)

## Discussion

### Summary of main findings

Many local authorities in England have introduced TMZs around schools, yet little is known about the perspectives of young people who are the most important target group of the policy. This qualitative study explored narratives of acceptability and perceived effectiveness of TMZs amongst young people attending secondary school in LAs with operating TMZs. We found that the young people were largely unaware of the policy both in general and in the context of their own school’s local food environment. They identified other aspects of the food environment as important contributors to poor diets and were open to further intervention in the school and broader food environment. The shortcomings of the school food environment and the widespread availability of unhealthy food and drinks from convenience and other stores were identified as significant barriers to the effectiveness of TMZs. They highlighted the important social role the food environment plays in their everyday lives and emphasised the need for alternative social infrastructure for socialising and passing time after school and on the weekends.

### Contributions to the literature

Our findings revealed that TMZs are generally acceptable to young people because they perceived that they do not change the existing food environment in a noticeable way or compromise their experience of it. Participant accounts revealed the influence of other aspects of the food environment on their diets, suggesting that takeaways are only one contributor to their consumption of unhealthy food and drinks. Previous research has highlighted the wide range of food environments through which adolescents can access unhealthy food and drinks in their everyday lives, including during school days, which often include travel through different spaces (Burningham & Venn, 2022; Caraher et al., 2014; Caraher et al., 2016; Cowburn et al., 2016; Tyrrell et al., 2017).

Consistent with previous studies, food outlets identified as popular with young people were heterogeneous and encompassed fast food outlets, takeaways, convenience stores, and supermarkets (Crawford et al., 2017; Wills et al., 2015). Access to social space, the friendliness of staff and the convenience, affordability and taste of the food and drink available were cited as the key factors affecting food-related decision-making. While proximity to food outlets was a consideration for younger participants, generally constrained to the school site at break and lunch times, older participants expressed a willingness to travel beyond the TMZ if their needs were not being met. This finding suggests a connection between break and lunchtime stay-on-site school policies, which are largely implemented in schools for students up to age 16 in England, and the potential of TMZs to impact dietary behaviours (Baines & Blatchford, 2019). With the freedom to leave site during break and lunch times and in their free study periods, the older participants in our study had more time to access foods in the external food environment than younger participants.

In contrast to previous research, which primarily focused on takeaways and fast food, participants reported most of their purchases as convenience food and drinks on account of their relative value for money and ready accessibility at all times of the day - before, during (if permitted to leave the school site) and after school (Forsyth et al., 2012; Patterson et al., 2012). The familiarity and social value of the branded products available, often heavily marketed to this demographic on digital platforms, were also described as important (Boyland et al., 2020; Buchanan et al., 2018; Stead et al., 2011).

The association between perceived shortcomings in the school food environment and patronage of external food outlets emerged as a key finding in our interviews. Participants frequently described long canteen queues and high prices as making the external food environment particularly enticing, and sometimes, necessary. The school food environment has been found elsewhere to act as “a push or pull factor” (Wills et al., 2015, p. x) with the external food environment, shaping young people’s food purchasing and consumption behaviours (Caraher et al., 2014; Caraher et al., 2016; Fletcher et al., 2014). The ‘takeaway experience’ was described at times as making up for the lack of commensal aspects of dining in school canteens. Participant accounts of struggling to have enough time to eat and socialise are consistent with recent research highlighting a significant reduction in the length of lunchtimes in UK secondary schools (Baines & Blatchford, 2019).

Consistent with previous research, our findings illustrate young people’s attachment to takeaway and fast food outlets as social spaces to gather with friends, ‘hang out’ and eat together (Burningham & Venn, 2022; Shaw et al., 2023; Thompson et al., 2018). Participants spoke of takeaway and fast food outlets as compensating for the lack of accessible and safe social infrastructure for young people, particularly in colder months (Webster, 2016). In line with this finding, a recent study into public acceptability of TMZs found a correlation between 16-17 year olds who perceive takeaways as important places for socialising *and* believe that young people would continue to patronise takeaways even if there were fewer near schools (Keeble et al., 2024). This further highlights the importance of takeaways in the social lives of young people and its related implications for policy acceptability.

Our findings suggest that changes in the digital food environment, particularly increased access to food delivery apps accelerated by the Covid-19 pandemic, may become a more significant barrier to TMZs in the future (Granheim et al., 2022; Kalbus et al., 2023). Although participants described schools as in some ways preventing access to FDAs, there was a sense that potential barriers could be circumvented. The schools in this study did not have specific policies on ordering food to the school site, and our research found no direct evidence that the food delivery aggregator companies ban deliveries to schools.

### Implications for policy and future research

Prior to the go-along interviews, participants in this study were generally unaware that TMZs were in operation around their school. Given the policy’s focus on restricting new takeaways, participants did not perceive any discernible changes in the external food environment. While recent evidence indicates that the policy has achieved its stated aim of reducing the growth of takeaways (Rahilly et al., 2024), participants seemed to conflate the lack of concrete change in the food environment with policy ineffectiveness.

Our research indicates that while TMZs may decrease the expected growth of takeaways in the longer term, they do not address other aspects of the food environment contributing to poor diets in young people and the wider population. TMZs would be more effective as part of a suite of policies designed to improve overall dietary health (Brown et al., 2021; Downs & Demmler, 2020). The ubiquitous presence of convenience foods and drinks around schools means that young people’s exposure to unhealthy foods is likely to persist, even with a potential reduction in takeaway outlets. This is also the case without greater regulation of unhealthy food marketing, particularly for products targeted at young people (Coleman et al., 2022; Neufeld et al., 2022).

As found in other studies, the participants in this study expressed a willingness to consume healthier options, yet generally struggled to source affordable healthier alternatives near their school (Calvert et al., 2020; Kelly et al., 2021). Participants frequently described their local food environments in critical terms that resonate with the concept of ‘food swamps’, where the high concentration of outlets selling unhealthy foods ostensibly ‘swamp out’ healthier alternatives (Fielding & Simon, 2011; Rose et al., 2011). Providing resource to schools or existing food outlets (identified as affordable to young people) to offer healthier alternatives or reformulate their menus may be a more effective short-term LA policy to encourage healthier eating.

Participants believed that issues with the school food environment were pushing students further toward making unhealthy choices in the external food environment. Enhancing the taste and affordability of school food, along with mitigating queues and allowing more time for socializing by extending lunchtimes, were identified as strategies to encourage students to opt for school-provided options. When schools are typically competing with the abundance of cheap convenience food readily available in the external environment, policies aimed at improving young people’s relationship with school food in ways that are qualitatively important to them are crucial. With reports indicating FDAs delivering to school premises in our study and others, schools may also be increasingly competing with unhealthy foods delivered directly into the school environment (Royal Society for Public Health, 2016). The regulation of FDAs to prohibit deliveries to schools could further support schools in cultivating a healthier food environment.

While the policy envisages a primary motivator in proximity, in reality the factors driving young people’s diet-related behaviours are more complex. For instance, although the policy may reduce exposure to takeaways in the long-term, it will not remove the secondary, and in some cases more important, need for safe and welcoming spaces for socialising. Given the social value of food environments, demonstrated in this study by the young people’s use of takeaway and fast food outlets as social spaces, interventions should also consider social and emotional needs (Hawkes et al., 2020; Isaacs et al., 2022).

### Strengths and limitations

This study generated unique data highlighting the perspectives of an important target population of TMZs: young people attending secondary school in England. The views of young people are not always considered in policy processes despite providing important and useful insights (Macauley et al., 2022; Rudner & Wilks, 2013). The observations and insights of young people from this study could be beneficial for LAs considering implementing TMZs around schools.

A strength of this study was the in-depth and rich data gained from go-along interviews. While participants may have been inhibited by the presence of the teacher/school chaperone, they were able to freely lead both the direction of the trip and the discussion, facilitating a dynamic exploration and fluid expression of the food environment in their own words. Including a heterogenous sample of young people in terms of age (11-18 years old), and socioeconomic background contributed to the development of themes reflecting the perspectives and experiences of secondary school pupils at different educational and developmental stages. However, despite our attempt to include equal numbers of male and female participants, including two single-sex schools resulted in a higher number of female participants. Furthermore, since our research is limited to the perspectives of secondary school pupils, future research into the perspectives of primary school pupils would allow for greater insight into the policy’s influence on dietary behaviours across different age groups.

This study is limited to urban areas of London and therefore is of limited applicability to other areas in the UK. While young people’s experiences of the food environment may vary in character and nature, their ideas on TMZs may be transferable to other contexts. Future research in more rural areas is needed to understand perceptions of TMZs in a wider range of social and physical contexts.

## Conclusions

This research has explored young people’s perspectives on TMZs around their schools and has in turn shed light on their wider experience and understanding of the food environment. While the focus on reducing the growth of takeaway outlets in the policy was acceptable amongst participants, their perception that there had been limited ‘change’ in the food environment decreased confidence in its ability to influence young people’s complex food behaviours. Interventions and policies should therefore consider the wide range of influences and motivators determining young people’s relationship with food. This involves considering the impact of unhealthy foods sold in food outlets other than takeaways and the influence of inadequate school food environments on young people’s interaction with the broader food environment. This study has also shown that young people are willing to engage in discussion about public health strategies and including engaging them in the policy process could increase acceptability and impact.

## Data Availability

All data produced in the present study are available upon reasonable request to the authors and in line with LSHTM ethics procedures.

## Acknowledgements

We express our gratitude to all the young people who generously contributed to this study, as well as the school staff who facilitated participant recruitment and the go-along interviews.

